# Mortality due to SARS COV-2 And its Associated Factors in East Shewa Zone Treatment Centers, Ethiopia, 2022: A retrospective cross-sectional study

**DOI:** 10.1101/2022.11.28.22282830

**Authors:** Jemal Hassen, Tewodros Getinet, Abera Botore, Mesfin Bekele, Bayissa Bekele, Firaol Jalata, Wake Abebe, Mekdes Sisay, Asnakech Getahun, Tadesse Ligidi

**Affiliations:** Oromia Health Bureau, Addis Ababa, Ethiopia; Adama Public Health Research & Referral Laboratory Center, P.O. Box 688, Adama, Oromia, Ethiopia; Saint Paul’s Hospital Millennium Medical College (SPHMMC) · School of Public Health

**Keywords:** SARS-CoV-2, Comorbidity, Ethiopia, Mortality, Laboratory test, Treatment Given

## Abstract

**Background:** Coronavirus disease (COVID-19) is an infectious disease that is caused by the SARS-CoV-2virus. The objective of this study was to determine SARS COV-2 Mortality and its associated factors in East Shewa Zone Treatment centers, Oromia, Ethiopia, 2022. The study of these types of viral infection will add new insight into the most common causes of mortality in SARS-CoV-2infection and the most common co-morbidities associated with the disease in the East Shewa Zone.

**Methods:** The study was conducted on patients who were admitted to Adama Hospital medical college and Modjo Primary Hospital for SARS-COV 2 treatment. Data used for the study were collected from March 2020-April 2022 GC. The study population was SARS-COV 2 patients who come to Adama Hospital and Medical College and Modjo Primary Hospital for treatment. All eligible SARS-CoV-2 patients’ data were collected from Both Adama and Modjo treatment center SARS-CoV-2 accession registration book and medical record card.

**Result:** A total of 409 patient data were collected from which 199 were from Adama Hospital and Medical College and 210 samples were collected from Modjo Primary Hospital Treatment center. The study design was a retrospective Cross-sectional study. The most affected age group in terms of mortality was the age group between 60-69 years old which suffers a 45.28% death rate. The major sign symptoms identified include cough (80.4%), Shortness of breath (66.7%) followed by fever (43.2%). SARS-CoV-2 Comorbidity was detected in 152 (37.2%) patients. Pneumonia was identified as the major comorbid disease to be recorded with 89(21.8%) cases. Other major comorbidities include Hypertension (16.9%) and Diabetes Mellites (13.9%). The least identified comorbidities were anemia (0.2%), Rectal cancer (0.2%), breast cancer (0.5%), and Chronic liver disease.

**Conclusion:** Nearly one in four (22.7%) SARS-COV 2 patients admitted for treatment to Adama Hospital and Medical College and Modjo Primary Hospital did not make their way out of treatment Hospitals alive. Pneumonia was identified as the major comorbid disease to be recorded with 89(21.8%) cases

## Introduction

### Background

Coronavirus disease (COVID-19) is an infectious disease that is caused by the SARS-CoV-2virus. The disease is caused by the novel coronavirus causing COVID-19. SARS-CoV-2 was first reported by officials in Wuhan City, China, in December 2019. All available evidence to date suggests that the virus has a natural animal origin and is not a manipulated or constructed virus. SARS-CoV-2believed to originated from a zoonotic source. It belongs to the subfamily Coronavirinae in the family Coronaviridae of the order Nidovirales and can cause respiratory, digestive, and nervous system diseases in humans and many other animals (1) (2) (3)(4) (5).

As an emerging acute respiratory infectious disease, COVID-19 primarily spreads through the respiratory tract, by droplets, respiratory secretions, and direct contact. In addition, it has been reported that SARS CoV-2 was isolated from fecal swabs and blood, indicating the possibility of multiple routes of transmission. However, this needs further clarification. The current data suggest an incubation period of 1–14 days, in most cases 3–7 days. The virus is highly transmissible in humans and causes severe problems, especially in the elderly and people with underlying chronic diseases. COVID-19 patients typically present with specific, similar symptoms, such as fever, malaise, and cough. Most adults or children infected with SARS-CoV-2have presented with mild flu-like symptoms, but a few patients are in critical condition and rapidly develop acute respiratory distress syndrome (ARDS), respiratory failure, multiple organ failures such as heart, brain, liver kidney, and even death (7)(8)(9).

COVID-19 is a systemic disease that can move beyond the lungs by blood-based dissemination to affect multiple organs. These organs include the kidney, liver, muscles, nervous system, and spleen. The primary cause of SARS-CoV-2mortality is acute respiratory distress syndrome initiated by epithelial infection and alveolar macrophage activation in the lungs (10). Another study in Iran reported older age (>65 years old), male gender (11) (12) hypertension, CVDs, diabetes, COPD and malignancies were associated with a greater risk of death from COVID-19 infection (11).

### Statement of the problem

Globally, as of 5:32 pm CEST, 12 October 2022, there have been 619,770,633 confirmed cases of COVID-19, including 6,539,058 deaths. Since the first cases were recognized in December 2019, SARS-CoV-2has spread fast around the world. Studies of Hospitalized patients have reported fatality rates ranging from 1.4% to 18.9%, and as high as 61.5% among those who were critically ill. There were also 6–10 Case fatality rates reported among older adults than among young children. Reported rates include 1.0% among adults aged 50–59 years, 3.5% among 60–69 years, 12.8% among 70–79 years, and 20.2% among 80 years or older (6).

### Significance of the study

The study of these types of viral infection will add new insight/knowledge into the most common causes of mortality in SARS-CoV-2infection and the most common co-morbidities associated with the disease. This study identified the most commonly reported health conditions that predispose individuals to SARS-CoV-2infection and subsequently lead to Hospitalization intensive care services and death in East Shewa Zone.

This type of study as of my knowledge, is not performed in East Shewa zone. So public health managers also may use the output of this study for planning for the management of this disease and the most commonly identified disease associated with SARS-CoV-2infection.

## Methodology

The objective of this study was to determine SARS COV-2 Mortality and its associated factors in East Shewa Zone Treatment centers, Oromia, Ethiopia, 2022.

### Study setting and study period

The study was conducted on patients who were admitted to Adama Hospital medical college and Modjo Primary Hospital. Adama Hospital and Medical College is one of the 1st Medical Hospital situated in Adama town, located in Oromia Region, 100 km to the southeast of Addis Ababa. Modjo Primary Hospital was founded in Modjo town which is 70 kilometers east of Addis Ababa. Based on the 2007 Census of the Central Statistical Agency of Ethiopia (CSA), this East Shewa zone has a total population of 1,356,342, of whom 696,350 are men and 659,992 are women. The total area of this zone is 8,370.90 square kilometers. East Shewa has a population density of 162.03. Adama Hospital Medical College and Modjo Primary Hospitals are public Hospitals assigned for SARS-CoV-2 treatment in the East Shewa zone (56).

### Source population

all SARS COV-2 cases in the East Shewa zone from SARS COV 2 treatment centers data collected between March 2020 and April 2022 GC. The study was conducted between April 2022 and Nov 2022.

### The study population

SARS-CoV-2 patients who come to Adama Hospital and Medical College and Modjo Primary Hospital SARS-CoV-2 for treatment centers data collected between March 2020 and April 2022 GC.

### Sample size and sampling technic

All SARS-CoV-2 patients’ data from March 2020 to April 2022 GC in Adama Hospital and Medical College (AHMC) and Modjo Primary Hospital (MPH) treatment centers were collected for analysis. A total of 409 patient data were collected from which 199 were from Adama Hospital and Medical College and 210 samples were collected from Modjo Primary Hospital Treatment center. Patients’ data were abstracted from Adama and Modjo treatment centers’ SARS-CoV-2 accession registration book and medical record card. Trained health professionals extract relevant data to pre-tested and standardized questionnaires. Missing variables were transferred from patients’ medical cards to the questionnaires. Patient medical cards and registration books are linked through unique identification numbers.

### Study design

Retrospective Cross-sectional.

### Source of data, data collection method, and tool

Source of data was treatment centers’ SARS-CoV-2 patient registration and medical registration card. Personnel working in these treatment centers carefully and systematically transfer data to the pretested questionnaire. Missing variable which cannot be completed either from patient registration or medical card will not be included in the study. Data clerks double-entered data into Epi Info 7.2.5.0 (Centers for Disease Control and Prevention, Atlanta). Data collector’s pre-test 10% of the questionnaire before actual data collection.

### Eligibility criteria

SARS COV2 patients data admitted to Adama Hospital Medical College (AHMC) and Modjo Primary Hospital (MPH) from March 2020 to April 2022 GC with complete data.

### Data quality

Health professionals working in AHMC AND MPH SARS-CoV-2 treatment centers who specifically participated in SARS-CoV-2 patient management were selected and given training on quality data collection. Data were double-entered in EPI INFO V.7 (Centers for Disease Control and Prevention, Atlanta), from questionnaires to minimize errors. At the end of each data collection day, the principal investigator reviewed each completed questionnaire.

### Data processing and analysis

Principal investigator checked the data for completeness and double entered into EPI INFO V.7 (Centers for Disease Control and Prevention). Data cleaning, coding, storing, and analysis were performed by using IBM SPSS V. 26 (Chicago). Descriptive analysis was conducted for each predictor variable. Each variable in the data underwent univariate analysis. Variables that scored p-value ≤ 0.25 in bivariate analysis were included in multiple logistic regression. In multiple logistic regression analysis, explanatory variables with p-value ≤ 0.05 were considered significant independent predictors for treatment outcome.

In multiple logistic regression analysis, the satisfactory of overall model fit was determined by the Omnibus test for model coefficients which was 0.000. For the model summary, we use Hosmer and Lemeshow Test which was 0.847 which indicates the overall model fitness was good.

### Variables of the study

#### Dependent variable

Treatment outcome of SARS-CoV-2 patients (Dead/Alive)

#### Independent variables

Age, sex, residence/address, sample collection date, case information (suspect, contact), symptoms present, types of sign symptoms, travel history, temperature, admission, patient outcome, lab test result, treatment taken

#### Ethical clearance

Oromia health bureau was responsible for issuing an ethical clearance. Data was held confidential and secure. Patient Identifiers were removed from variables during analysis.

For the sake of anonymity, Authors other than principal investigators were not given access to patient data identifiers.

#### Dissemination and Utilization of Results

Oromia health bureau received the output finding. The result of the investigation was communicated to all relevant bodies. The research team provides the result of the investigation to AHMC and Modjo primary Hospital. The manuscript will be submitted to a peer-reviewed journal.

## Result

### Selection of study participants

Refer for Diagram for steps in selection of study participants in the study of SARS COV-2 Mortalities And its Associated Factors in East Shewa Zone Treatment Centers, Oromia, Ethiopia, 2022 in the figure document.

#### Patient demographics

A nearly equal number of study participants from AHMC and MPH were enrolled in the study. The proportion includes 51.3% from MPH and the rest of the participants were from AHMC. The majority of the patients presented to this treatment center were male (63%). Nearly 80% of study participants were aged between 18-65 of which only 1.7% were under 18 years old. Most of the study participant’s residence districts were from other towns outside Adama and Modjo. Most (64.1%) study participants do not have a travel history and in 70.9% of the patient’s families. (Table 1)

The most affected age group in terms of mortality was the age group between 60-69 years old which suffers a 45.28% death rate. Death was not registered in the age category of <10 years and between 10-19 years old patients. (Figure 1)

**Figure 1.**
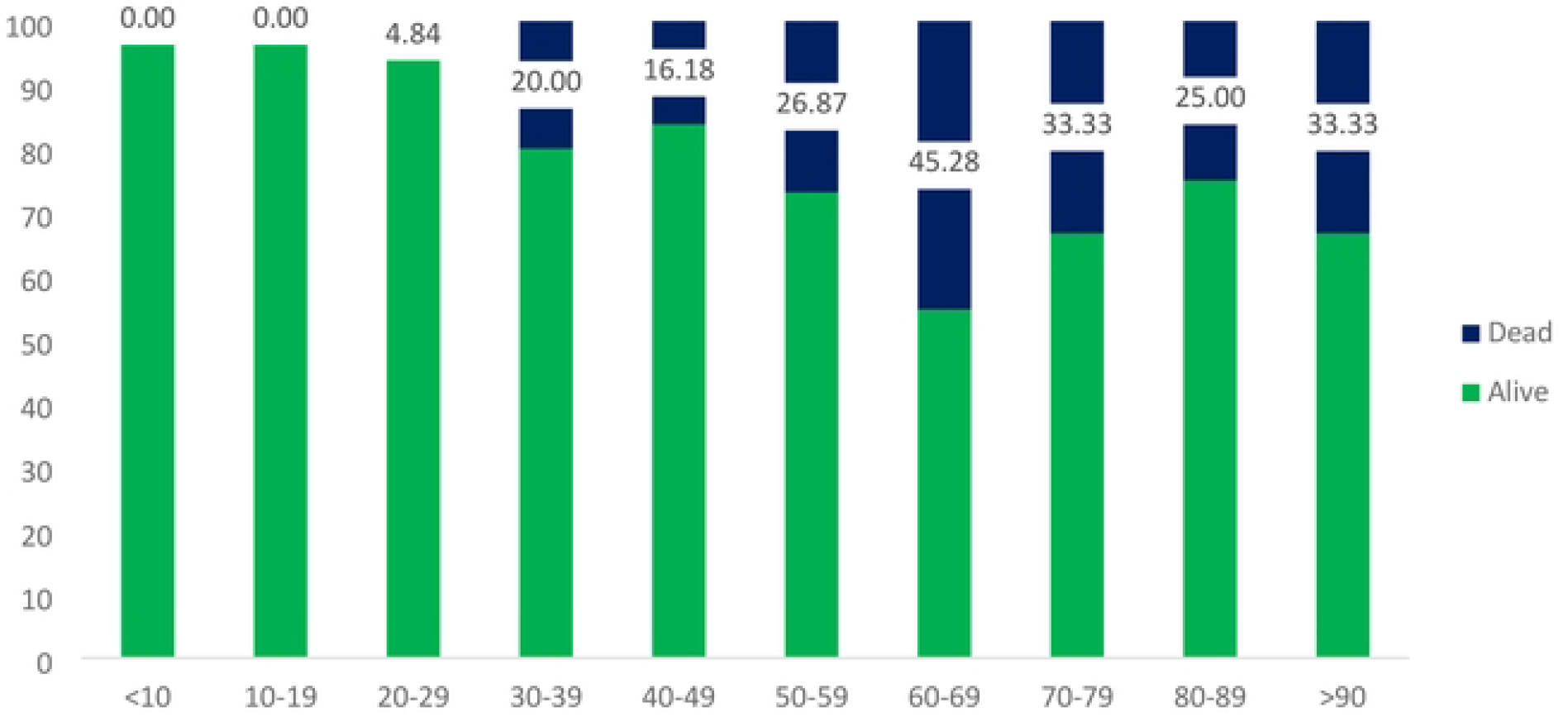
The proportion of Dead Patients in different age categories with SARS COV 2 disease in the study of SARS COV-2 Mortalities And its Associated Factors in East Shewa Zone Treatment Centers, Oromia, Ethiopia, 2022.

**Figure 1.**
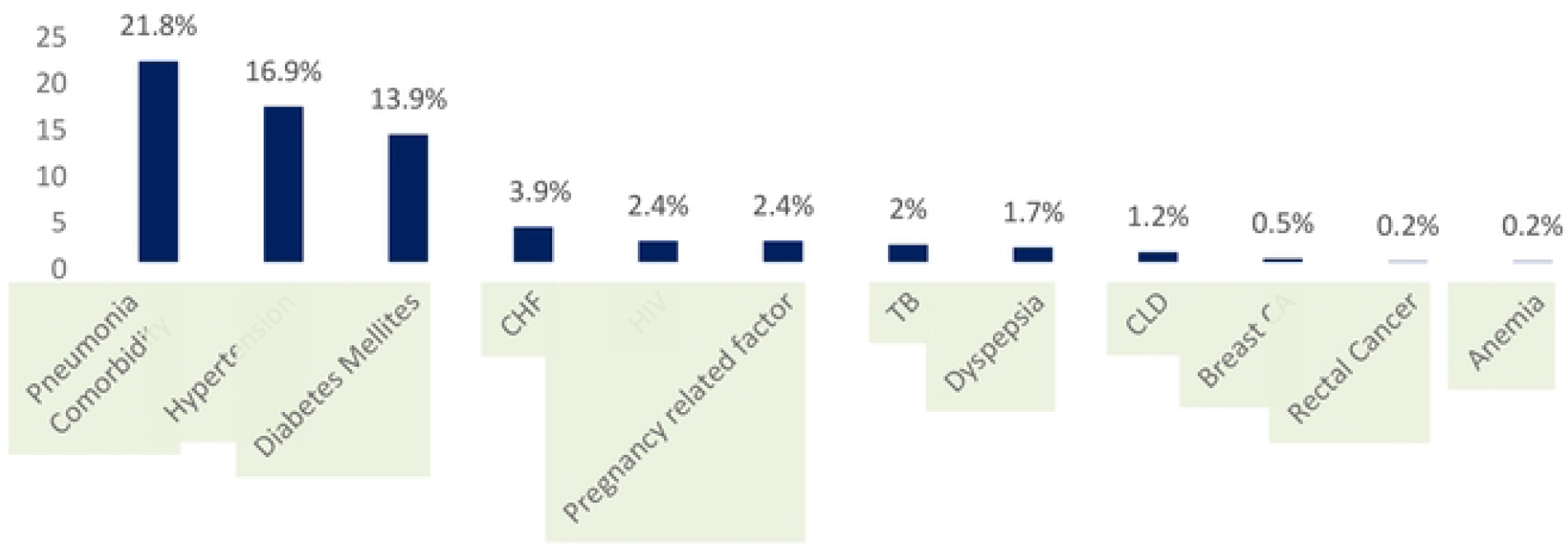
Cornorbidities in the study of SARS COV-2 Mortalities And its Associated Factors in East Shewa Zone Treatment Centers, Oromia, Ethiopia, 2022.

### Sign symptoms in SARS-CoV-2 patients in East Shewa zone

The study identified a multitude of signs and symptoms in SARS-CoV-2 patients. The major sign symptoms recorded include cough (80.4%), Shortness of breath (66.7%) followed by fever (43.2%). Convulsion and Orthopnea were observed in only one case each. Other sign symptoms which were least observed in this study include Epistasis (0.49%), Pitting edema (0.49%), Joint pain (2.20%), Loss of smell and test (3.18%), Asthma (3.42%), Vomiting (4.16%) and Sore throat (4.65). (Table 2)

The study identified comorbidity in 152 (37.2%) patients. Pneumonia was identified as the major comorbid disease to be recorded with 89(21.8%) cases. Other major comorbidities include Hypertension (16.9%) and Diabetes Mellites (13.9%). The list identified comorbidities were anemia (0.2%), Rectal cancer (0.2%), breast cancer (0.5%) and Chronic liver disease (CLD).

Identified complications include Acute kidney injury (2%) and acute respiratory distress syndrome (1.7%) were recorded. (Table 3)

The mean Hospital stay in days of SARS-CoV-2 patients in the East Shewa zone treatment center was found to be 13.5 days. The minimum number of Hospital stays was one day. The maximum, standard deviation and range of 61 days, 10.0 days, and 60 days respectively. (Figure 3)

**Figure 2.**
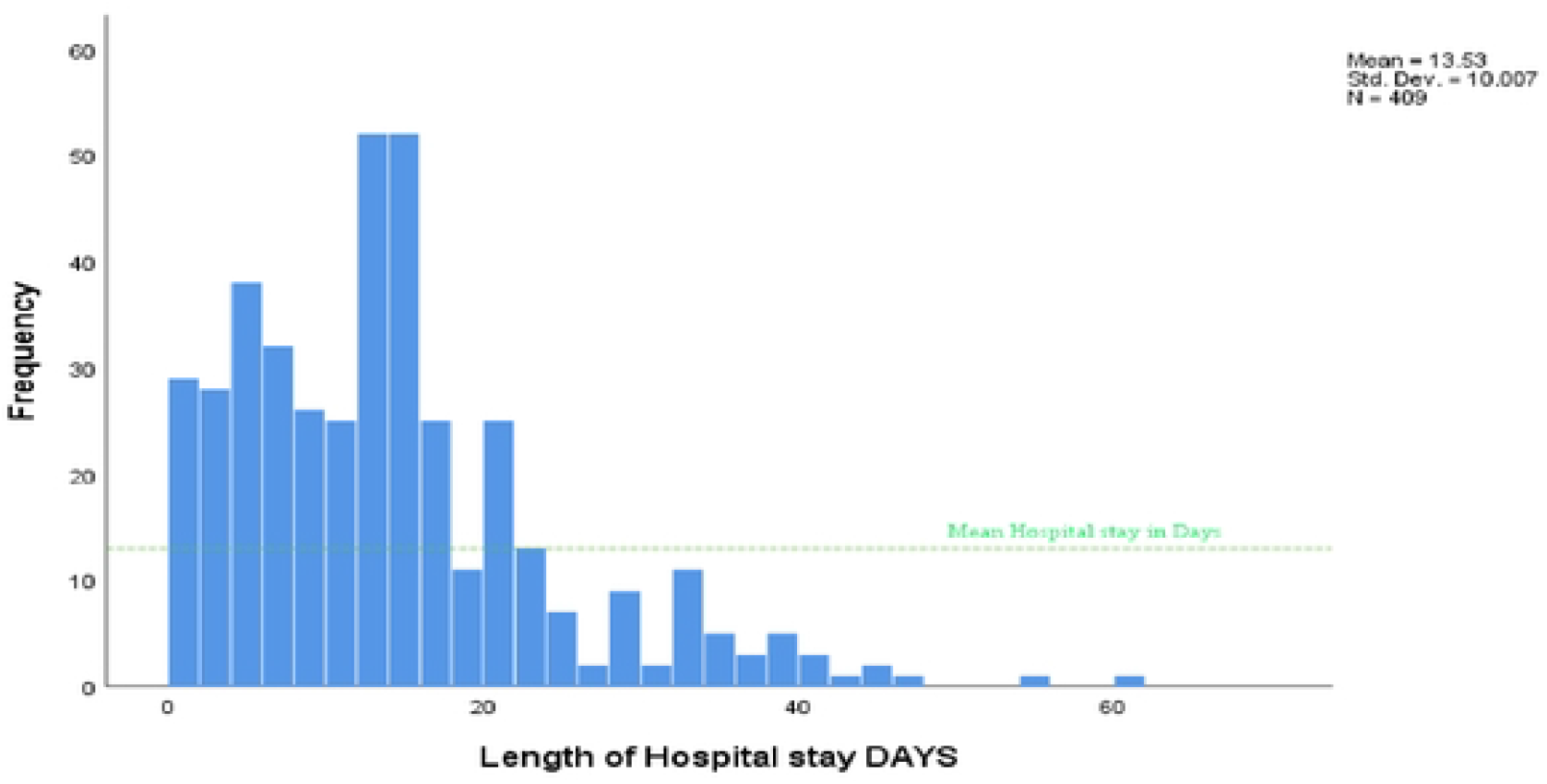
Length of Hospital stay in days in the study of SARS COV-2 Mortalities And its Associated Factors in East Shewa Zone Treatment Centers, Oromia, Ethiopia, 2022.

**Figure 3.**
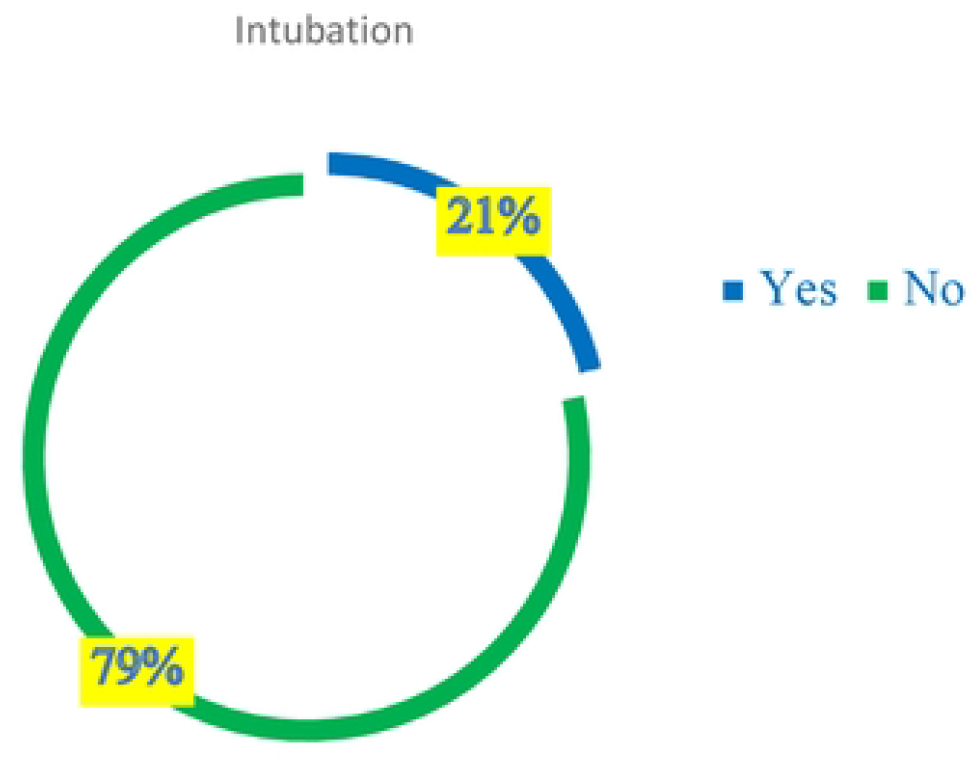
Proportion of patients receiving Intubation procedure in East Shewa zone SARS COV-2 treatment Centers in the study of SARS COV 2 Mortalities And its Associated Factors in East Shewa Zone Treatinent Centers, Oromia, Ethiopia, 2022.

The investigator also studied SARS-CoV-2 disease severity. The proportion of patients with severe disease was found to be 13.0%. 18.3%. of SARS-CoV-2 patients were found to have mild SARS-CoV-2 disease. (Table 4)

SARS-COV-2 patients requiring intubation were found to be 1.3%. (Figure 4). Nearly one in four (22.7%) SARS-CoV-2 patients reported to Adama Hospital and Medical College and Modjo Primary Hospital did not make their way out of SARC COV 2 treatment centers in East Shewa zone alive. (Figure 5)

**Figure 4.**
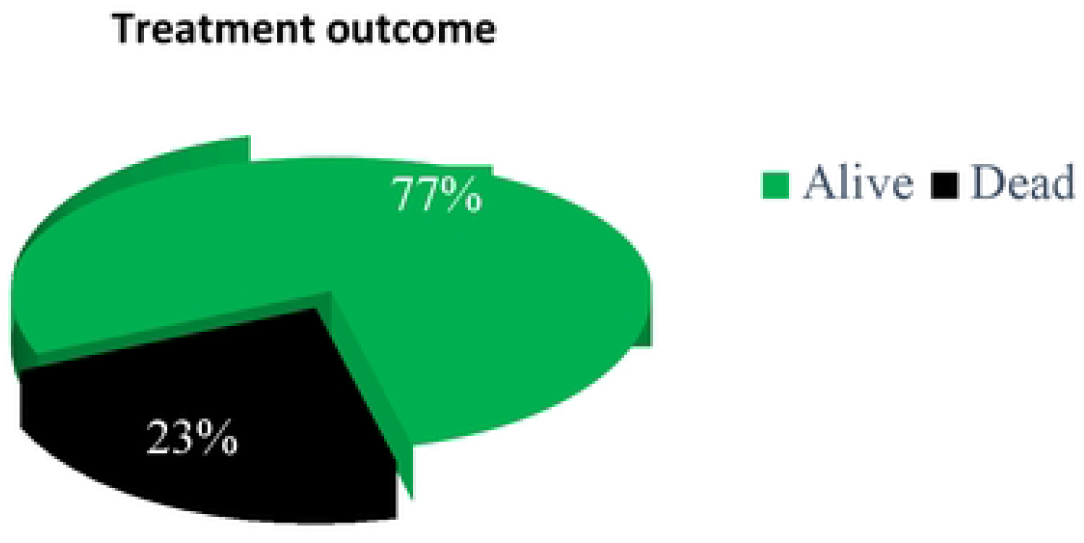
Treatment outcome in the study of SARS COV-2 Mortalities And its Associated Factors in East Shewa Zone Treatment Centers, Oromia, Ethiopia, 2022.

The most prescribed drug type in the East Shewa zone during the study period for SARS-CoV-2 treatment was Dexamethasone with well more than half of patients receiving it. SARS-CoV-2 treatment centers prescribed Dexamethasone (66.6%), Vancomycin (57%), Medical Oxygen (55%) and Unfractionated Heparin (UHF) (43.5%). The table below summarizes the types and frequency of treatment types given during the study period in East Shewa zoneSARS-CoV-2 treatment centers. (Table 5)

Physicians at AHMC and MPH treatment centers in the East Shewa zone ordered different laboratory tests. The most widely ordered laboratory test was Serum Creatinine (26.6%) of patients followed by blood urea with 24.9%. (Table 6)

In this study, the most affected age group was the 18-65 years age group which accounts for about 79.5% of SARS-CoV-2 patients. This age group is also the most affected in terms of mortality. About 72.0% of all death fromSARS-CoV-2 treatment center was from this age group. Male accounts for about 66.7% of death compared to female SARS-CoV-2 patients. Among the study participants, 40.9% of patients who are dead were found to have at least one type of comorbidity. Among intubated SARS-CoV-2 patients 88.2% were found to be deceased. (Table 7)

In analyzing data using Binary logistic regression, the Goodness of fit test was checked by using Hosmer and Lemeshow Test. This value was found to be 0.652 which is greater than the 0.05% cutoff value which fails to reject the null hypothesis that states no factors do not affect treatment outcome in SARS-CoV-2 infection. The model summary also was found to be good in explaining the variance in the treatment outcome of SARS-CoV-2 infection. The Pseudo R square (Nagelkerke R Square) test was found to be 0.881. This indicates that our predictor variable was able to explain 88.1% of the variance in treatment outcome in SARS-CoV-2 infection.

In bivariate analysis, variables with a p-value of ≤ 0.25 were directly used for analysis in multiple logistic regression. Age (p-value 0.01), Residence (p-value 0.00), presence of other cases in the family (p-value 0.124), cough (p-value 0.25), fever (p-value 0.08), headache (p-value 0.06), SOB (p-value 0.02), Easy fatigability (p-value 0.09), Sore throat (p-value 0.09), Muscle pain (p-value 0.18), CHF (p-value 0.15), Dyspepsia (p-value 0.04), LOS (p-value 0.00), ARDS (p-value 0.04), and Intubation (p-value 0.00), were directly inserted into multiple logistic models using enter method.

In multiple logistic regression, variables with p-value ≤ 0.05 were selected as a significant factor in affecting treatment outcomes in the East Shewa zone treatment center. Variables with p-value ≤ 0.05 but that cross value 1 will not be included in a set of significant factors for treatment outcome. In this study residency district of being in Adama (p-value 0.01, CI: 1.71-5.60), LOS < 7 days (p-value 0.01, CI: 9.88-42.63) being intubated (p-value 0.00, CI: 17.96-25.24) were found to be a significant factor that affects treatment outcome in East Shewa zoneSARS-CoV-2 infection.

Those SARS-CoV-2 patients who lived in Adama town are 9.8 times more prone to the occurrence of death than patients who lived in Modjo town. SARS-CoV-2 patients with a length of stay of < 7 days in Hospital in East Shewa treatment centers are 205 times riskier than patients with LOS > 21 days in Hospital. In this study, SARS-CoV-2 patients with ARDS are 9.78 times more prone to death than patients without ARDS complications.

## Discussion

The study of treatment outcomes of SARS-CoV-2 patients attending Adama Hospital Medical College and Modjo Primary Hospital (which are Government Hospitals selected as SARS-CoV-2 treatment centers by Oromia regional government) was conducted from March 2020 to April 2022 GC. A One-year-old child SARS-CoV-2 patient was the minimum age to be recorded in treatment centers and a 92-year-old patient was the maximum age of patient receiving treatment. The mean and median age of the study participant was 47 years old. This age group is much lower than the median age group in a study participant of SARS COV-2 comorbidity network and outcome in Hospitalized patients in Crema, Italy by Tommaso Gili (40) in 2021, in which the median age group was 70.8 years of age. This difference could be accounted for by the population age distribution difference between the two countries.

This study finds that male patients accounted for 63% of Hospitalization due to SARS-CoV-2 infection. The case fatality rate (CFR) among male SARS-COV-2 patients in the study area was 62%. This finding was similar to the rate of Hospitalization of SARS-CoV-2 patients in Australia (2) in which the rate of Hospitalization by male patients was between 54.3% and 68%. This indicates the probability that sex does not have much variation across geographical differences and socio-economical inequalities.

In this study, we identified a range of signs and symptoms of SARS-CoV-2infection. These signs and symptoms include Cough, fever and SOB. Muscle pain, chest pain, vomiting, Loss of smell and taste, headache and sore throat. These sign and symptoms finding were similar with finding in a study by Richard H. Hunt et al (23), Jorge A. Huete-Perez et al (57), Laura McArthur (2) and Stephan Ludwig et al. (3). Considerable difference was observed across different research findings in different countries including this study regarding the prevalence of sign and symptoms of SARS-CoV-2 infection. In our study, the prevalence of cough, SOB and fever during SARS-CoV-2 infection was 80.4%, 66.7% and 43.2% respectively. But in the study of Richard H. Hunt et al the prevalence of cough, SOB and fever were found to be 68.9%, 71.2% and 71.6% respectively. The prevalence of cough was found to be higher in this study than compared to the study by Richard H. Hunt et al, Jorge A. Huete-Perez et al,and Laura McArthur et al. study.

In the study of comorbidity of SARS-CoV-2 infection in East Shewa zone SARS-CoV-2 treatment centers, Pneumonia, Hypertension, Diabetes Mellites, Rectal cancer, Breast cancer, Chronic liver disease, coronary heart disease (CHD), Human Immuno-Deficiency Syndrome (HIV), Pregnancy related disease, Tuberculosis (TB), Dyspepsia and Anemia were identified as comorbidities of SARS-CoV-2. This finding of comorbidities was similar to the study conducted by Negin Ebrahimi et al, Sumit Jamwal et al, and Jorge A. Huete-Perez in a different study setting. In our study, the prevalence of hypertension was 16.9%. This finding was similar to the finding of the prevalence of hypertension by Gypsyamber D’Souza (47), Soo Lim (22) and Mei Peng (41), which is 18.6%, 17.1% and 15-22%, But the prevalence of hypertension in our study was different from the following studies, Study by author Priya V which is 21.1%, Nazar Zaki (33) (53%), Anamika Gupta (35) which is 64.6% and Irawaty Djaharuddin (58) which was 42%. The difference in the prevalence of hypertension in this study area may be due to different socio-economic attributes such as lifestyle and economic status of the respective community.

There was a similarity and difference in the prevalence of Diabetes Mellitus (DM) in patients with SARS-CoV-2 patients in our study and other studies. The prevalence of DM in our study was found to be 13.9%. This finding was much lower than the prevalence of DM in the SARS-CoV-2 patients in the study of Soo Lim et al (22) 33-58%, Anamika Gupta (35) 28.1% Irawaty Djaharuddin (58) 28.2 and slightly lower prevalence than a study by Carmine Savoia (27) 19% and Jose-Manuel Ramos-Rincon et al (34) 17.4%, But slightly higher than the study by Priya Veluswamy (14) in which the prevalence of DM in SARS-CoV-2 patients was 9.7%. This difference could be attributed to the difference in the socioeconomic status of the study settings.

Different research papers were produced indicating the case fatality rate of SARS-CoV-2 infection across the world. In our study, the case fatality rate across all sex and age categories was found to be 23% (6). This prevalence rate was higher that the study by Chris R. Triggle et al in Qatar in which the overall case fatality rate of the disease was found to be 8%. This difference could be attributed to the difference in case management of SARS-CoV-2 patients in the Hospitals and the availability of anti-retroviral treatments in Qatar which may lead to the increased survivability of patients in Qatar. In another study of the case fatality rate of SARS-CoV-2 patients by Tommaso Gili et al (40), the rate was found to be 17.5% which is slightly lower than that of our study. This difference could also be attributed to the difference in the availability of better treatment options in Italian Hospitals.

The case fatality rate of this disease was also studied based on age category. According to this study, we found that the minimum case fatality rate was observed in the age category of <10 years old and the age category between 10-19 years of age. This finding was similar to the study done by Margarita V. Revzin (18), which founds that child was less affected by SARS-CoV-2 disease and the case fatality rate among children was minimum. The maximum case fatality rate was observed in the age category between 60-69 years of age. But in a study conducted by Tommaso Gili (40), the maximum case fatality rate was observed in the age category > 90 years of age. In a study conducted by Tommaso Gili (40), the case fatality rate in the age group 60-69 years of age was reported to be 6.6%. In our study Case fatality rate among the age group, 80-89 years of age was found to be 25%. This finding was similar to research findings done by Mohan Kumar Muthu Karuppan in the USA which found the case fatality rate among the same age group was found to be 10-27%.

## Data Availability

Data set used in this study are available upon request from editorial manager of other relevant body upon request

## Conclusion and recommendation

The most affected age group in terms of mortality was the age group between 60-69 years old which suffers a 45.28% death rate. Death was not registered in the age category of <10 years and between 10-19 years old patients. Major sign symptoms recorded include cough (80.4%), Shortness of breath (66.7%) followed by fever (43.2%). Convulsion and Orthopnea were observed only in one patient in each category. Other sign symptoms which were least observed in this study include Epistasis (0.49%), Pitting edema (0.49%), Joint pain (2.20%), Loss of smell and test (3.18%), Asthma (3.42%), Vomiting (4.16%) and Sore throat (4.65). The least of comorbidities in patients with SARS-CoV-2 disease in East Shewa zone SARS-CoV-2 treatment centers. In this study, comorbidity was detected in 152 (37.2%) patients. Pneumonia was identified as the major comorbid disease to be recorded with 89(21.8%) cases. Other major comorbidities include Hypertension (16.9%) and Diabetes Mellites (13.9%). The least identified comorbidities were anemia (0.2%), Rectal cancer (0.2%), breast cancer (0.5%) and Chronic liver disease (CLD).

Since the majority of death occurred in older age, the government is better to introduce care for the older age population group. The government also needs to avail treatment for Pneumonia Hypertension and DM disease since these diseases were found to be the major comorbidity in his study area. We recommend also better management of non-communicable diseases in the East Shewa Zone.

## Declarations

### limitations of the study

Government archive/ secondary data was used for the analysis of mortality of SARS-COV-2 in East Shewa zone treatment centers. Data were collected for other purposes. This may make it difficult to frame variables that are essential to study the mortality of SARS-COV-2 in the study area.

### Ethics approval and consent to participate

Ethical Approval was obtained from the Oromia regional health bureau, Adama Public Health Research and Referral Laboratory Center IRB board which is legally responsible to issue such approval. Oromia regional health bureau, Adama Public Health Research and Referral Laboratory Center IRB board provides us with a local language version of the institutional review board letter of approval.

### Consent for publication

I am the principal investigator. The funding institution gave me full responsibility to manage this research starting from proposal development to publication, dissemination and publication in a reputable peer-reviewed journal.

### Availability of data and materials

Data used for this study is available in the principal investigators’ data archive and funding organization archive. Data can be readily available upon request from the responsible bodies including requests from publishers.

### Authors’ contributions

- JH: Principal investigator, Corresponding author*
- AB: responsible for organizing, reviewing and summarizing literature
- TG: Inferential statistic specialist and interpreted final result,
- MB: Overall coordinate the study
- BB: Reviewed planning/proposal of the study
- FJ: Curator
- WA: Reviewed descriptive statistics
- MS: Reviewed Discussion, Conclusion and recommendation part of the study
- AG: Data collector and software specialist
- TL: Infographics analysis

## Acknowledgments

I would like to extend my sincere appreciation to Adama Public Health Research and Referral Laboratory Center for allowing me to conduct this study in which this project would not be possible without the financial and managerial support of this study. I also would like to thank Adama Hospital Medical College and Modjo Primary Hospital management and clinical staff for their dedicated cooperation towards achieving this study. My special appreciation and thanks go to my previous Biostatistics lecturer at Saint Paulo’s Hospital and Medical College for his guidance on statistical concepts. Finally, I would like to thank Adama Public Health Research and Referral Laboratory Center staff for their valuable input in the finalization of this study.

## Abbreviations

AC: Anticoagulant
AHMC: Adama Hospital Medical College
AKI: Acute Kidney Injury
ALP: Alkaline phosphatase
ALT: Alanine transaminase
APHRRLC: Adama Public Health Research and Referral laboratory Center
ARDS: Acute Respiratory Distress Syndrome
AST: Aspartate Aminotransferase
BSC: Bachelor of Science
BUN: Blood Urea Nitrogen
Ca: Calcium
CA: Cancer
CDC: Center For Disease Control and Prevention, USA
CHF: Cardiac Heart Failure
CLD: Chronic Liver Disease
CLD: Chronic Liver Disease
CNS: Central Nervous System
COPD: Chronic obstructive pulmonary disease
COVID 19: Coronavirus 19
CSA: Central statistics Agency, Ethiopia
CT: computerized tomography
CVD: Cardio Vascular Disease
CXR: Chest X ray
DM: Diabetes Mellitus
EC: Ethiopian Calendar
EPIINFO: Epidemiological Information
HIV: Human Immuno deficiency Virus
HTN: Hypertension
IBM SPSS: International Business Machines, Statistical Package for Social Sciences
ICU: Intensive Care Unit
IQR: Inter quartile Range
IRB: Institutional review board
K: Potassium
LOS: Length of Stay
MPH: Masters of Public Health
Na: Sodium
NYC: New York City
P VALUE: Probability value
SARS: COV2 Severe Acute Respiratory Syndrome - Corona Virus - 2 sCr Serum Creatinine
SGOT: serum glutamic-oxaloacetic transaminase
SGPT: Serum Glutamic Pyruvic Transaminase
SOB: Shortness of breath
TB: Tuberculosis
UHF: Unfractionated Heparin
UK: United Kingdom
USA: United States of America
VBM: Variant Being Monitored
VOC: Variant of Concern
VOHS: Variant of High Consequence
VOI: Variant of Interest
WBC: White Blood Cell
WHO: World Health Organization

---

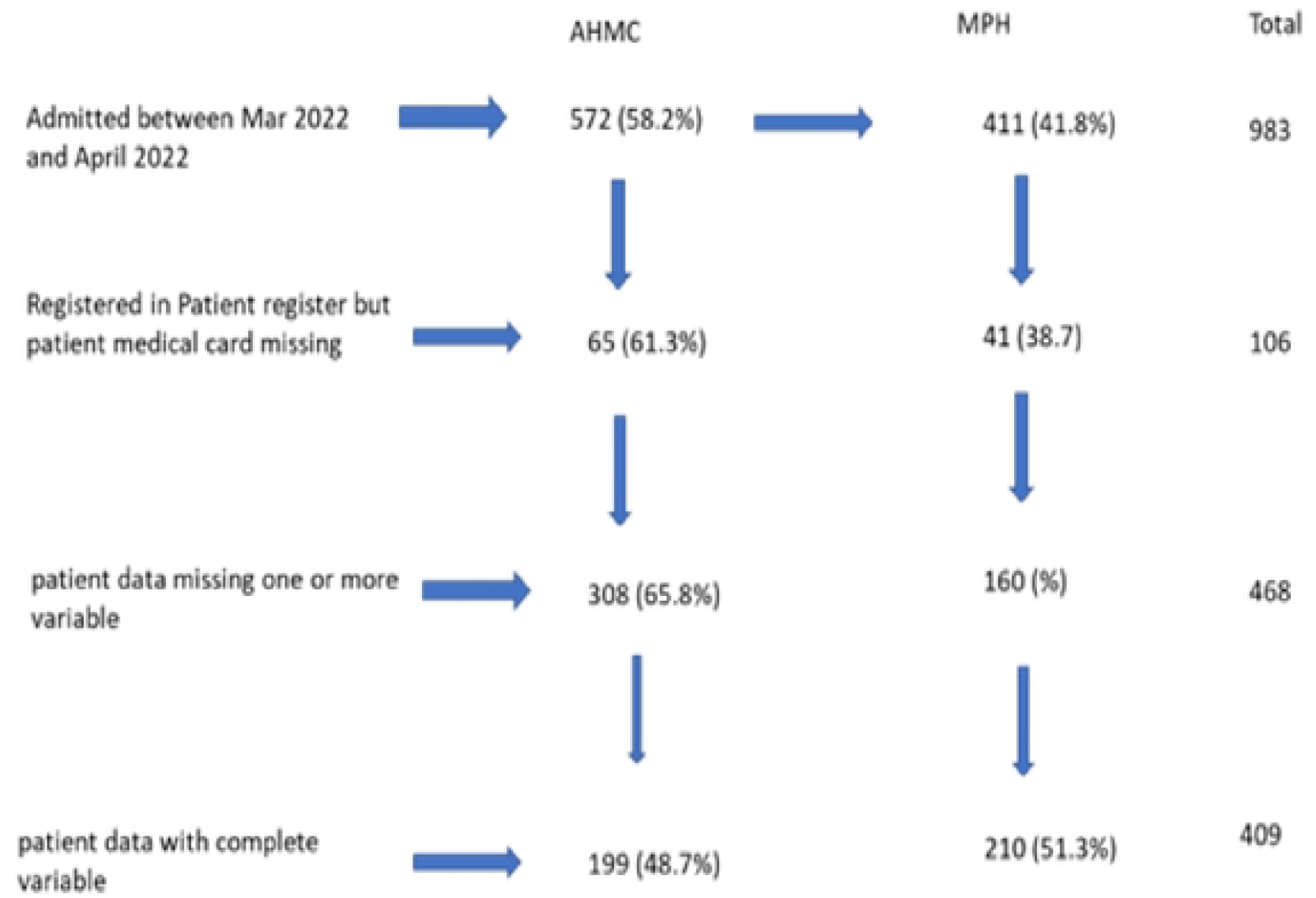

Diagram for selection of study participants in the study of SARS COV-2 Mortalities And its Associated Factors in East Shewa Zone Treatment Centers, Oromia, Ethiopia, 2022.

